# Inflammatory biomarker signatures in post-surgical drain fluid may detect anastomotic leaks within 48 hours of colorectal resection

**DOI:** 10.1101/2022.08.05.22278403

**Authors:** Simone M. Cuff, Nicola Reeves, Eifion Lewis, Eleri Jones, Sarah Baker, Athanasios Karategos, Rachel Morris, Jared Torkington, Matthias Eberl

## Abstract

**Background:** The optimal treatment of colorectal cancer is surgical resection and primary anastomosis. Anastomotic leak can affect up to 20% of patients and creates significant morbidity and mortality, a leak is based on clinical suspicion and subsequent radiology. Peritoneal biomarkers have shown diagnostic utility in other medical conditions and could be useful in providing earlier diagnosis. This pilot study was designed to assess the practical utility of peritoneal biomarkers after abdominal surgery utilising an automated immunoassay system in routine use for quantifying cytokines.

**Method:** Patients undergoing an anterior resection for a rectal cancer diagnosis were recruited. A peritoneal drain was placed in the proximity of the anastomosis during surgery, and peritoneal fluid was collected at day 1 and 2 post-operatively, and analysed using the Siemens IMMULITE platform for IL-1β, IL-6, IL-10, CXCL8, TNF-α and CRP.

**Results:** 42 patients were recruited. Anastomotic leak was detected in 4 patients and a further 5 patients had other intra-abdominal complications. The IMMULITE platform was able to provide robust and reliable results from the analysis of the peritoneal fluid. A metric based on the combination of peritoneal IL-6 and CRP levels was able to accurately diagnose three anastomotic leaks, whilst correctly classifying all negative control patients including those with other complications.

**Conclusion:** This pilot study has demonstrated that a simple immune signature in surgical drain fluid could accurately diagnose an anastomotic leak at 48 hours post-operatively using instrumentation that is already widely available in diagnostic laboratories.

## Introduction

Colorectal cancer is the third most common cancer worldwide, with the majority of patients treated with surgical resection and primary anastomosis. Between 1 and 20% of colorectal resections result in an anastomotic leak (AL) ^1^. AL poses a significant risk for patients. As well as the immediate threat of developing faecal peritonitis and sepsis, with its attendant morbidity and mortality profile, in the long term AL is associated with a lower five year cancer-specific survival rates ^2–7^ and increased risk of cancer recurrence ^2–4,8,9^. Hence, there is a need to identify AL quickly to enable early intervention, possible anastomotic salvage and avoid long term complications.

Current methods of diagnosing AL are usually based on serial clinical examinations and radiological imaging from days 3-5 post-operatively. These include symptoms such as pain, tachycardia, fever, oliguria, ileus, diarrhoea and leukocytosis, in combination with elevated blood levels of C-reactive protein (CRP). However, the clinical signs are non-specific and are only observed once there is a systemic response to the AL. Similarly, blood CRP can aid diagnosis, with post-operative day 3 levels of less than 172 mg/l having a negative predictive value of 97% but only a low specificity for diagnosis of an AL ^10^.

By post-operative day 3 to 5 the risk of mortality increases, alongside the necessity for an emergency operation to treat faecal peritonitis with its concomitant high risk of a permanent stoma formation. Earlier detection of an AL could change the management and treatment of these patients and, crucially, improve their post-operative outcome and quality of life.

In the present study, we examined whether peritoneal fluid drained from the surgical site can be used to diagnose AL more rapidly and accurately than current practice. By assaying peritoneal fluid in the proximity to the anastomotic site, the local response to colonic contamination of the peritoneum should be detectable more quickly, sensitively, and specifically than later systemic responses. This hypothesis was based on previous studies in which peritoneal fluid was found to respond more rapidly to the presence of AL than routinely sampled blood ^11^, confirming that it is more likely to be an effective medium for robust early detection. Similarly, our own research in different pathologies has shown that local immune signatures (“immune fingerprints”) at inflammatory sites are powerful predictors of acute infections, including in the peritoneal cavity of patients receiving peritoneal dialysis ^12,13^, in the urine of patients presenting with suspected urinary tract infection ^14^, and in cerebrospinal fluid of neurosurgical patients ^15^. In order to have immediate relevance for patient benefit, the present study was designed to assess the practical utility of biomarkers in drain fluid after abdominal surgery and test a suite of clinically approved biomarkers using the Siemens IMMULITE platform, an automated immunoassay system routinely used for quantifying cytokines in hospitals globally.

## Methods

### Ethics approval

All methods were carried out in accordance with relevant guidelines and regulations. Experimental protocols were approved by Cardiff University, and written informed consent was obtained from all subjects. Recruitment of patients for this study was approved by the Wales Cancer Bank under reference no. 18/016 and conducted according to the principles expressed in the Declaration of Helsinki.

### Objectives

The primary objective of the study was to assess whether altered levels of peritoneal fluid biomarkers (cytokines) in response to AL could be measured on a widely available commercial immunoanalyser platform.

The secondary outcome was the analysis of peritoneal biomarker measurements to explore their sensitivity and specificity for AL diagnosis.

### Patient cohort

Between June 2019 and June 2021, all patients undergoing an elective anterior resection within the colorectal department of the University Hospital of Wales were screened for inclusion. Inclusion criteria were cancer diagnosis, intended primary anastomosis, age ≥18 years and capacity to consent. Exclusion criteria were non-cancer diagnosis, lack of capacity and a permanent end colostomy formation. Patients who had neoadjuvant chemoradiotherapy were included, and the intention to form a defunctioning ileostomy was not an exclusion criterion. Operations were performed according to surgeon preference, either laparoscopically or via a midline laparotomy. Laparoscopic procedures had extraction sites via a midline or a Pfannenstiel incision. The primary anastomosis was formed using a conventional circular stapler, the size according to surgeon choice. At the end of the operation a non-suction drain was placed in the left iliac fossa, into the pelvis.

### Clinical assessments

Patient demographics were collected pre-operatively. These included age, sex, BMI, smoking status, and previous chemotherapy and/or radiotherapy. Intraoperative variables were also collected, including operation time, laparoscopic or open approach, and formation of defunctioning stoma. Post-operatively, data were collected on length of hospital stay, tumour staging (TNM) and post-operative complications. Complications were classified as AL or other complications, which included bleeding, small bowel obstruction and any other reason for return to theatre. AL was defined as clinically manifest insufficiency of the anastomosis leading to a clinical state requiring treatment (i.e. grade B/C)^16^. AL was confirmed by either CT scan and/or reoperation. As such patients were stratified into three groups for analysis – ‘Uneventful Recovery’, ‘Anastomotic Leak’ and ‘Other Complications’. All clinical data were incorporated into a database in which data were pseudo-anonymised by assigning each patient a study number. No personally identifiable information was included.

### Collection and Storage of Peritoneal Fluid

Drain peritoneal fluid was collected at 8:00 am on days 1, 2 and 3 post-operatively. The first sample (day 1) was collected between 14 to 20 hours post-operatively. All samples were transferred for analysis in the laboratory within 30 minutes. The peritoneal fluid was centrifuged twice at 500 *g* for 20 minutes at 20°C temperature. The cell-free supernatant was removed, aliquoted into 2 ml cryotubes and stored at −80°C until further use.

### Peritoneal drain fluid analysis

Peritoneal fluid levels of interleukin (IL)-1β, IL-6, IL-8/CXCL8, IL-10, tumour necrosis factor (TNF)-α and C-reactive protein (CRP) were measured using the corresponding IMMULITE 1000 kits on IMMULITE 1000 Immunoassay systems through the solid-phase chemiluminescent immunometric assay (CLIA) method. All kits and instruments were supplied by Siemens Healthineers GmbH, Germany. Normal assay range; IL-1β: 5-1000 pg/ml; IL-6: 2-1000 pg/ml; CXCL8: 5-7500 pg/ml; IL-10: 5-1000 pg/ml; TNF-α: 4-1000 pg/ml; and CRP: 0.3-100 mg/l. For CRP measurements, samples were prediluted 1 in 100 in a CRP-free protein/buffer matrix prior to assay. Due to inherent high concentrations of IL-6 in peritoneal fluid, two dilutions (1 in 50 and 1 in 100) were used to cover the reasonable expectations of dilutional requirements, using an IL-6-free protein/buffer matrix prior to assay. All other biomarkers were measured directly in the undiluted fluid.

### IMMULITE assay performance

To evaluate precision, single donor or pooled peritoneal fluid samples were selected and prepared at different concentrations, divided into aliquots, and stored at −20°C until use, with a fresh aliquot used for each run. For IL-1β, IL-6 and IL-10, repeatability/within lab precision, the prepared pools were run once per day, for 5 days, in replicates of 5. For CRP, TNF-α and CXCL8, repeatability was assessed from 10 replicates run from the same sample aliquot in a single run. Linearity was assessed by diluting a peritoneal fluid pool with an analyte concentration near the upper reportable limit (or as high as the sample pool would allow), with a peritoneal fluid pool containing low concentrations of the relevant analyte to yield concentrations of 100%, 75%, 50%, 25%, 10%, 5%, 2.5%, 1%, and 0% of the original high-concentration pool. Separate pools were prepared for each analyte. All dilutions were tested in duplicate. Linearity of IMMULITE assays for peritoneal drain fluid was then analysed by weighted regression. To evaluate recovery of cytokines, peritoneal fluid samples were spiked with 2-3 solutions containing different amounts of each respective analyte. Samples for determination of recovery were prepared by making stock solutions in analyte-free protein buffer matrix. Stock serum solutions were added to peritoneal fluid samples at a ratio of 1:20. Spiked peritoneal fluid samples were assayed in duplicate, and the ratios between observed and expected concentrations were calculated.

### Data analysis

Data were analysed in R4.1 (R core team, 2017)^17^ and graphed in Prism 9 (GraphPad Software, San Diego, California USA). Biomarker thresholds were determined by maximising F1 scores across all timepoints. The F1 score is the harmonic mean between precision and recall, and is optimal for datasets with a large proportion of true negative patients. To create biomarker algorithms, log2 concentrations in pg/ml (CRP) or ng/ml (IL-6) were combined into an unweighted linear metric, the 6C metric, with the boundary optimised by F1 score. Biomarker weighting was tested but found to not improve accuracy.

## Results

### Patient demographics

The study cohort comprised a total of 42 patients who underwent an anterior resection between June 2019 and June 2021. Patient demographics did not differ significantly between groups (**Table 1**). In particular, there was no difference in operation time or the prevalence of laparoscopic versus open approach to operating between the three groups.

**Table 1.** Patient demographics and characteristics

|  |  | Anastomotic<br>leaks (*n*=4) | Other<br>Complications<br>(*n*=5) | Uncomplicated<br>recoveries (*n*=33) |
| --- | --- | --- | --- | --- |
| **Age in years, mean (range)** |  | 56 (31-82) | 61 (54-70) | 66 (40-87) |
| **Sex** | Male | 3 | 2 | 17 |
|  | Female | 1 | 3 | 16 |
| **BMI, mean (range)** |  | 32.1 (21.3-41.0) | 27.1 (23.4-37.0) | 27.5 (19.3-35.3) |
| **Smoking status** | Smoker | 1 | 0 | 6 |
|  | Non-Smoker | 3 | 2 | 21 |
|  | Ex | 0 | 3 | 6 |
| **Chemotherapy** | Yes | 1 | 1 | 3 |
|  | No | 3 | 4 | 30 |
| **Radiotherapy (Y:N)** | Yes | 1 | 1 | 5 |
|  | No | 3 | 4 | 28 |
| **Approach** | Laparoscopic | 3 | 4 | 27 |
|  | Open | 1 | 1 | 6 |
| **Operation time in minutes,**<br>**mean (range)** |  | 285 (210-390) | 225 (140-320) | 250 (120-435) |
| **Defunctioning**<br>**ileostomy** | Yes | 1 | 4 | 13<br>(1 other*) |
|  | No | 3 | 1 | 19 |
| **Tumour stage**<br>**T1:T2:T3:T4a** | T1 | 0 | 1 | 4<br>(2 other**) |
|  | T2 | 0 | 2 | 6 |
|  | T3 | 4 | 2 | 16 |
|  | T4 | 0 | 0 | 5 |

\* Hartmann's procedure. \*\* one patient no histological presence of tumour cells (previous radiotherapy), one patient T0 (pre-operative biopsies of high grade dysplasia suggestive of adenocarcinoma but no presence of adenocarcinoma histologically post operatively).

There were 9 patients who had post-operative complications; 4 patients had an AL and 5 had other complications including 3 patients with small bowel obstruction requiring re-operation and 2 patients with significant post-operative bleeds. 33 patients showed uncomplicated recoveries. Patients who experienced post-surgical complications stayed on average 7 days longer in hospital than patients with no such complications. Hospital length of stay for patients with AL was increased even further (**Figure 1**), demonstrating the cost of AL to both patient health, and to health providers.

**Figure 1.**
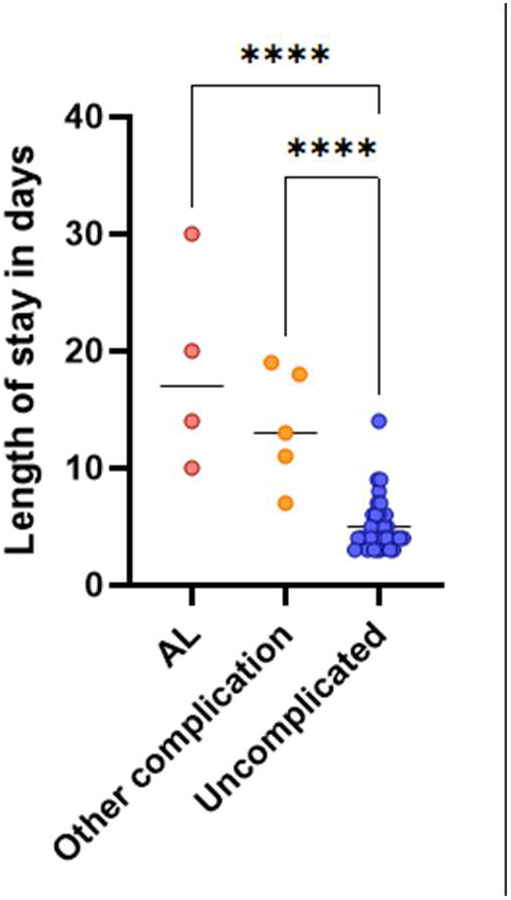
Length of hospital stay for patients undergoing colorectal surgery with uncomplicated recoveries compared to those with anastomotic leaks (AL) or other complications. Each data point indicates an individual patient; horizontal lines display the group mean. Indicated groups were significantly different by one-way ANOVA with Tukey’s post-hoc test for multiple comparisons. ****, *p*<0.0001.

### Validation of drain fluid as matrix for the IMMULITE platform

The IMMULITE 1000 platform is currently only approved for plasma and serum measurements. Therefore, we first determined whether peritoneal drain fluid was a suitable substrate for the chosen measurement system. Results were found to be robust and repeatable, with coefficients of variance between 2.2% and 10.3% (**Table 2**) showing that surgical site drain fluid is an excellent matrix for determining local cytokine levels using this platform. This was confirmed by assessing linearity and range of detection against ELISA-based detection, and spike-recovery profiles (**Table 3**). Together, these data demonstrate that the IMMULITE 1000 platform is capable of consistently determining cytokine concentrations in surgical site drain fluid to clinically relevant levels of precision.

**Table 2.**
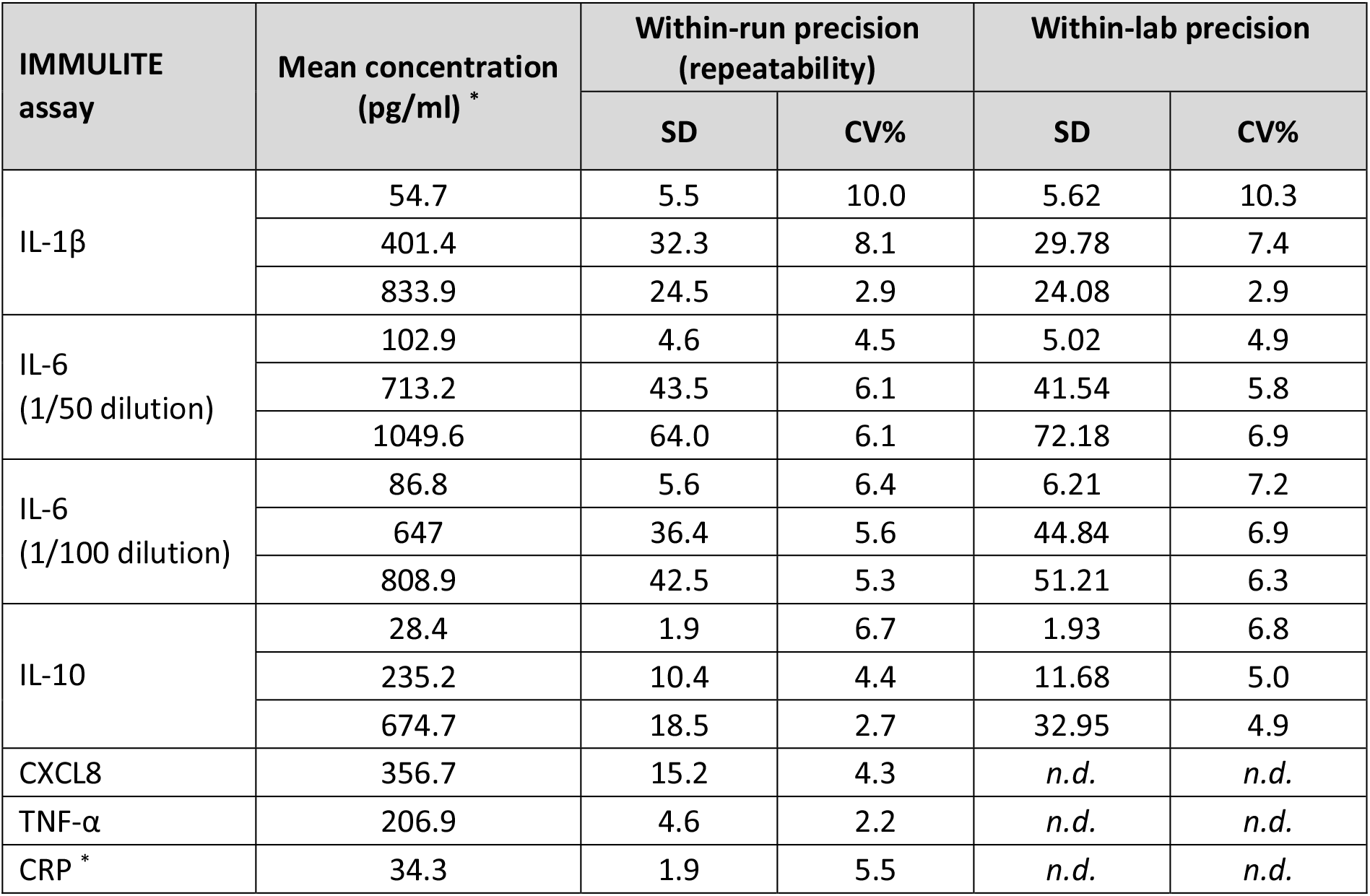
IMMULITE assay precision. Within-run and within-lab precision to measure IL-1β, IL-6 and IL-10 was determined using the 5×5 design of a minimum of 5 test days (1 run per day) with 5 replicate measurements for each sample per run. Within-run precision to measure CRP, CXCL8 and TNF-α was determined from a single sample, single run, in 10 replicates each. SD, standard deviation; CV, coefficient of variation. *n*.*d*., not done. CRP was measured in mg/L.

**Table 3.** IMMULITE assay linearity and precision. Linearity was analysed by weighted regression. All dilutions were tested in duplicate. To determine recoveries, spiked samples were assayed in duplicate, and observed versus expected doses were calculated.

| IMMULITE assay | Linearity | Recovery |
| --- | --- | --- |
| IL-1β | 13.35-875.7 pg/ml | 92-101% |
| IL-6 (1/50 dilution) | 24.69-1002.8 pg/ml | 104-110% |
| IL-6 (1/100 dilution) | 12.84-948.6 pg/ml | 106-109% |
| IL-10 | 28.35-666.9 pg/ml | 99-107% |
| CXCL8 | 31-228 pg/ml | 92-109% |
| TNF-α | 268-3600 pg/ml | 113-114% |
| CRP | 0.1-107 mg/L | 103% |

### Peritoneal immune signatures after colorectal surgery

Analysis of the peritoneal drain fluid on the IMMULITE platforms demonstrated measurable amounts of the inflammatory biomarkers IL-1β, IL-6, IL-10, CXCL8, TNF-α and CRP (**Figure 2**). Diagnostic accuracy was high for all biomarkers, partly reflecting the fact that all chosen biomarker thresholds correctly classified 93-97% of unaffected patients (**Table 4**: NPV). Given that this comprises the vast majority of patients, diagnostic accuracy was correspondingly high across the entire cohort independent of the ability to correctly identify the small number of AL cases (**Table 4**: PPV). F1 score instead does not use true negatives in its calculation and so is more suitable for situations in which there is an imbalance between group sizes. Here, amongst individual biomarkers, F1 score was highest for IL-6. This could be increased further by creating a log2-based derivative algorithm which gave equal weights to IL-6 and CRP readings (the 6C metric; **Figure 3**). Indeed, the 6C metric only misclassified a single AL patient. This is particularly impressive given that IL-6 and CRP would also be expected to increase in other (non-infectious) scenarios of inflammation. In addition, the 6C metric correctly classified all negative control patients as well as five patients with non-AL complications.

**Table 4.** Discrimination between patients with AL and all other patients at day 2 post-operation by individual markers and derived metrics. Scores above 0.95 and optimal *p* value are highlighted in bold italics. Threshold values for positive/negative: IL-1β, 320 pg/ml; IL-6, 75 ng/ml; IL-10, 300 pg/ml; CXCL8, 5900 pg/ml; CRP, 70 pg/ml; TNF-α, 400 pg/ml; 6C combined IL-6/CRP algorithm, 5.8. PPV, positive predictive value; NPV, negative predictive value. *Fisher’s Exact Test.

| | IL-1β | IL-6 | IL-10 | CXCL8 | CRP | TNF-α | 6C Metric |
| --- | --- | --- | --- | --- | --- | --- | --- |
| Sensitivity | 0.75 | 0.75 | 0.50 | 0.50 | 0.50 | 0.50 | 0.75 |
| Specificity | 0.79 | **0.96** | **0.96** | 0.86 | **1.0** | 0.93 | **1.0** |
| PPV | 0.33 | 0.60 | 0.67 | 0.33 | **1.0** | 0.50 | **1.0** |
| NPV | **0.96** | **0.96** | 0.93 | 0.93 | 0.94 | 0.93 | **0.97** |
| Diagnostic accuracy | 0.79 | 0.94 | 0.91 | 0.82 | 0.94 | 0.88 | **0.97** |
| F1 score | 0.46 | 0.75 | 0.57 | 0.40 | 0.67 | 0.50 | 0.86 |
| Significance (*p*)* | 0.052 | 0.0031 | 0.035 | 0.14 | 0.011 | 0.059 | **0.00081** |

**Figure 2.**
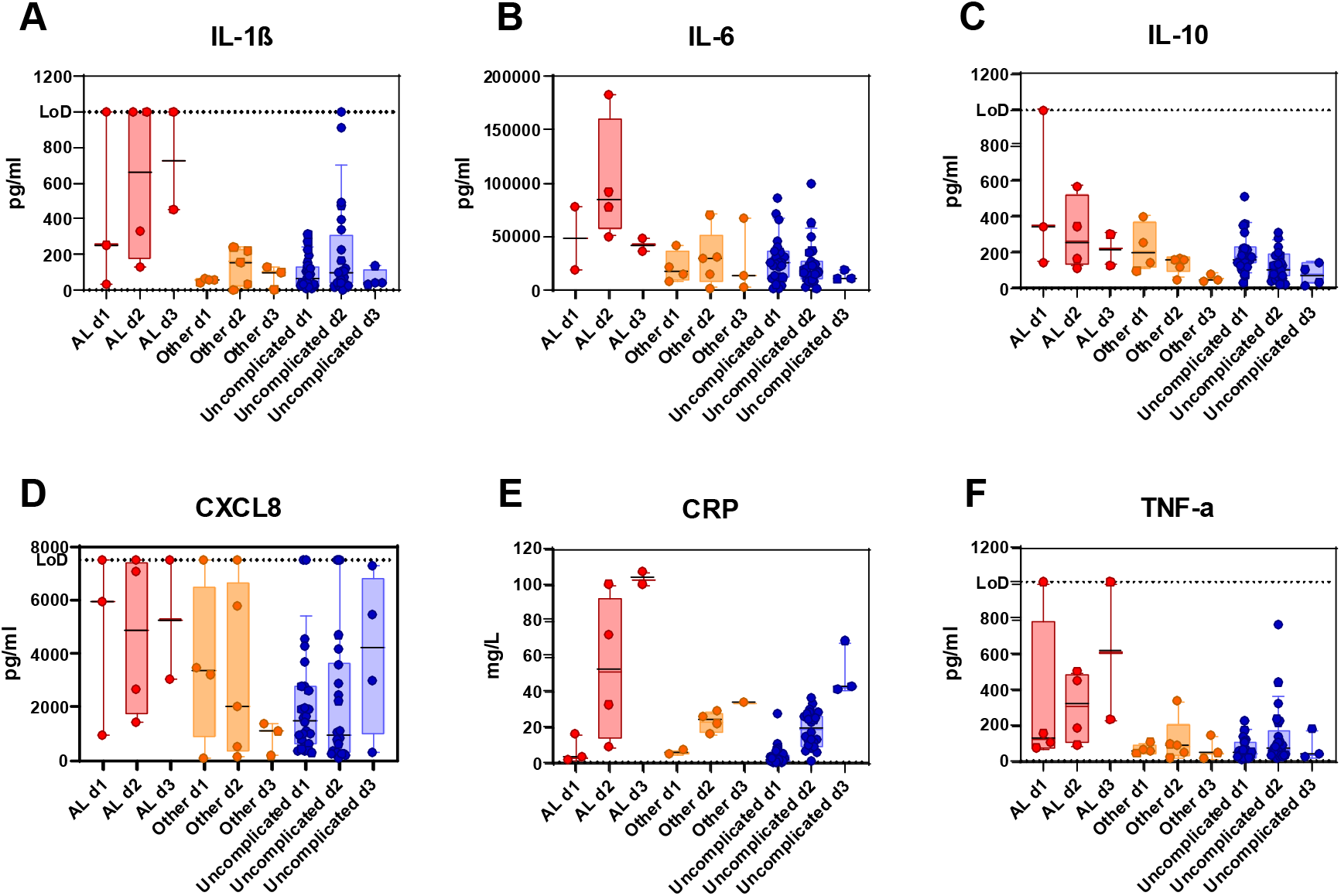
Cytokine levels in the drain fluid of patients with AL (red), other complications (orange), or uncomplicated surgeries (blue) over the first three days post-surgery. Each data point indicates an individual patient; boxes show interquartile range and whiskers extend from the 10^th^ to 90^th^ centiles. LoD, Limit of detection.

**Figure 3.**
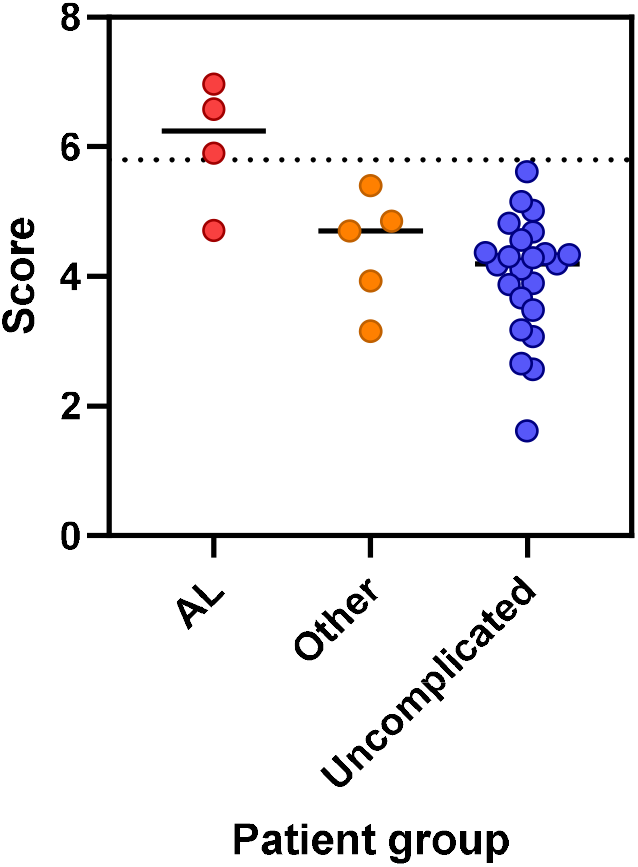
Discrimination between AL patients and all other patients at day 2 after surgery, using an algorithm based on local levels of IL-6 and CRP. Each data point indicates an individual patient; horizontal lines show group mean. Dotted line indicates the threshold between negative and positive values.

## Discussion

For patients undergoing a surgical colorectal resection, AL is a significant complication that changes the post-operative pathway, with increased short-term morbidity and mortality, and long-term reduced cancer specific survival. Analysis of the peritoneal fluid of the patients included in this study has demonstrated that inflammatory biomarkers are both measurable on a commercial platform and could be accurate in diagnosing an AL in the early post-operative period.

All biomarkers tested, with the exception of CXCL8, showed a measurable increased level in peritoneal fluid in those patients with AL at day 3, in agreement with previous published studies ^11,18^. IL-6 is an inflammatory cytokine which is upregulated in response to microbial pathogens but also tissue injury. Consequently, when measured in blood it is a non-specific marker of systemic inflammation ^19^. Peritoneal IL-6 has been measured before in AL studies and has been proposed as predictive marker for the presence of AL in patients after surgery in isolation ^20^. It is broadly successful, with sensitivities and specificities in line with our measurements, but is insufficiently accurate to be used in isolation. In contrast, the present study shows that it can be combined specifically with local CRP levels derived from drain fluid to create an effective metric that can detect AL within 48 hours post-operatively with high positive and negative predictive values.

CRP is commonly measured in blood where it is non-specifically increased in inflammatory and infective conditions, including in post-operative patients with no complications. CRP can act as an anti-bacterial effector which binds surface polysaccharides, opsonising bacteria including those that would be released into the peritoneum when an AL occurs, and activating the classical complement cascade. Systemic levels of CRP can be used as a negative predictive value of AL between 3 and 5 days post-operatively ^10,21^, in keeping with its responsiveness to inflammation. CRP is commonly characterised as an acute phase protein made by hepatic cells in response to IL-6 and IL-1β ^22^, however production can also be stimulated in macrophages ^23^. Our detection of elevated CRP in drain fluid 48 hours post-operatively support the hypothesis that it is produced locally in response to leakage of colonic content. Importantly, the differences in drain fluid CRP levels between AL patients and those experiencing inflammation due to ot her complications at this time point show that measuring local CRP has the potential to be a more specific biomarker for AL than systemic CRP levels.

Dual detection of blood levels of IL-6 and CRP has already been proposed as biomarkers of AL ^24^. However, while both markers have excellent negative predictive values, leading to high AUROC scores, positive predictive values for AL have remained poor so far. Our current study shows that sampling of drain fluid has both greater sensitivity for AL detection than blood markers and gives reliable results 24 hours earlier. The magnitude and early detection of the inflammatory response likely reflect the proximity of the fluid to the site of the inflammatory event, which is in agreement with studies that have shown IL-6 release from peritoneal membranes within 2 hours of surgery ^25^. Interestingly, IL-10, which we found at levels averaging 200 times lower than IL-6, was below the level of detection in the same study. The use of drain fluid rather than blood could also potentially mitigate the general nature of IL-6 and CRP being increased in response to unrelated inflammatory stimuli, such as concurrent infections and systemic conditions. This has been shown to be the case in other contexts such as meningitis, where it was found that blood IL-6 was not as specific a marker as IL-6 in the cerebrospinal fluid ^15^.

While promising, this is a pilot study and needs to be replicated with more samples in a larger study. Given the distribution of the readings, it is possible that in a larger cohort IL-10 or IL-1β may also prove to be useful in a combined metric with IL-6, and investigating whether these could add to the robustness of the metric will be an important next step to the research. The patient cohort measured in the present study was also limited to those undergoing an anterior resection, and results were not stratified on tumour location in relation to the peritoneal fold or the distance of tumour/anastomosis from the anal verge. Without the inclusion of all pathologies (for example cancer and inflammatory bowel disease) and all colonic resections, it is difficult to assess the applicability of the results to all anastomosis and patients. However, this is the first study to indicate a sensitive and specific marker for AL before clinical manifestation.

The impact of an earlier diagnosis of an AL is hard to predict at present, and further research is needed into the best management strategy for patients who are predicted to develop an AL. However, an accurate method to diagnoe and thus the ability to intervene and reduce the need for a defunctioning ileostomy, with its associated morbidity and need for reversal, is likely to ameliorate the risk of AL-related complications before the development of systemic sepsis and improve the patient’s pathway.

## Conclusion

This pilot study shows that is possible to measure peritoneal biomarkers in surgical drain fluid and utilise a combined metric to detect AL within 48 hours of an anterior resection, using instrumentation that is widely available in large hospitals. The rapid detection of AL will allow further research into early interventions, which should improve patient outcomes including reduction in length of stay, reduced mortality and morbidity and long term cancer survival.

## Data Availability

All data produced in the present study are available upon reasonable request to the authors.

## Acknowledgements

We are grateful to all patients for participating in this study, and to the clinicians and nurses for their cooperation. We would also like to thank Corinne Nguyen and Ian Weeks for their support and advice throughout this project, and the Wales Cancer Bank for facilitating patient recruitment and sample collection.

This study was not preregistered as it was an exploratory pilot study, examining the ability to detect peritoneal biomarkers in both the laboratory and commercial platform. Data will be available on reasonable email request to the corresponding author.

## Notes

Funding Sources This research was supported by the European Regional Development Fund via the Welsh Government Accelerate programme, a Wellcome Trust Institutional Translational Partnership Award (ITPA), a Cancer Research Wales “Access to Patient Samples for Translational Research Award” (ASTRA), and the Wales Data Nation Accelerator (WDNA) scheme.

Conflict of Interest Statement EL, EJ and RM are employees of Siemens Healthineers. Neither the funders nor Siemens Healthineers had any impact on the study design, data collection, data analysis, decision to publish or preparation of this manuscript.

### Competing Interest Statement

EL, EJ and RM are employees of Siemens Healthineers. Neither the funders nor Siemens Healthineers had any impact on the study design, data collection, data analysis, decision to publish or preparation of this manuscript.

### Funding Statement

This research was supported by the European Regional Development Fund via the Welsh Government Accelerate programme, a Wellcome Trust Institutional Translational Partnership Award (ITPA), a Cancer Research Wales Access to Patient Samples for Translational Research Award (ASTRA), and the Wales Data Nation Accelerator (WDNA) scheme.

### Author Declarations

Recruitment of patients for this study was approved by the Wales Cancer Biobank under reference no. 18/016 and conducted according to the principles expressed in the Declaration of Helsinki. The Wales Cancer Biobank is licensed by the Human Tissue Authority under the UK Human Tissue Act (2004) to store human tissue for research (licence no. 12107) and is approved as a Research Tissue Bank by Wales Research Ethics Committee (REC) 3.

## References

1. McDermott FD, Heeney A, Kelly ME, Steele RJ, Carlson GL, Winter DC. Systematic review of preoperative, intraoperative and postoperative risk factors for colorectal anastomotic leaks. Br J Surg. 2015;102(5):462–79.

2. Fujita S, Teramoto T, Watanabe M, Kodaira S, Kitajima M. Anastomotic leakage after colorectal cancer surgery: a risk factor for recurrence and poor prognosis. Jpn J Clin Oncol. 1993;23(5):299–302.

3. Petersen S, Freitag M, Hellmich G, Ludwig K. Anastomotic leakage: Impact on local recurrence and survival in surgery of colorectal cancer. Int J Colorectal Dis. 1998;13(4):160–3.

4. Merkel S, Wang WY, Schmidt O, Dworak O, Wittekind CH, Hohenberger W, et al. Locoregional recurrence in patients with anastomotic leakage after anterior resection for rectal carcinoma. Color Dis. 2001;3(3):154–60.

5. Walker KG, Bell SW, Rickard MJFX, Mehanna D, Dent OF, Chapuis PH, et al. Anastomotic leakage is predictive of diminished survival after potentially curative resection for colorectal cancer. Ann Surg. 2004;240(2):255–9.

6. McArdle CS, McMillan DC, Hole DJ. Impact of anastomotic leakage on long-term survival of patients undergoing curative resection for colorectal cancer. Br J Surg. 2005;92(9):1150–4.

7. Lin JK, Yueh TC, Chang SC, Lin CC, Lan YT, Wang HS, et al. The Influence of Fecal Diversion and Anastomotic Leakage on Survival after Resection of Rectal Cancer. J Gastrointest Surg. 2011;15(12):2251–61.

8. Akyol AM, Mcgregor JR, Galloway DJ, Murray GD, George WD. Anastomotic leaks in colorectal cancer surgery: a risk factor for recurrence? Color Dis. 1991;6(4):179–83.

9. Bell SW, Walker KG, Rickard MJFX, Sinclair G, Dent OF, Chapuis PH, et al. Anastomotic leakage after curative anterior resection results in a higher prevalence of local recurrence. Br J Surg. 2003;90(10):1261–6.

10. Singh PP, Zeng ISL, Srinivasa S, Lemanu DP, Connolly AB, Hill AG. Systematic review and meta-analysis of use of serum C-reactive protein levels to predict anastomotic leak after colorectal surgery. Br J Surg. 2014;101(4):339–46.

11. Reeves N, Vogel I, Ghoroghi A, Ansell J, Cornish J, Torkington J. Peritoneal cytokines as a predictor of colorectal anastomotic leaks on postoperative day 1: a systematic review and meta-analysis. Tech Coloproctol [Internet]. 2022;26(2):117–25. Available from: https://doi.org/10.1007/s10151-021-02548-y

12. Zhang J, Friberg IM, Kift-Morgan A, Parekh G, Morgan MP, Liuzzi AR, et al. Machine-learning algorithms define pathogen-specific local immune fingerprints in peritoneal dialysis patients with bacterial infections. Kidney Int. 2017;92(1):179–91.

13. Goodlad C, George S, Sandoval S, Mepham S, Parekh G, Eberl M, et al. Measurement of innate immune response biomarkers in peritoneal dialysis effluent using a rapid diagnostic point-of-care device as a diagnostic indicator of peritonitis. Kidney Int. 2020;97(6):1253–9.

14. Gadalla AAH, Friberg IM, Kift-Morgan A, Zhang J, Eberl M, Topley N, et al. Identification of clinical and urine biomarkers for uncomplicated urinary tract infection using machine learning algorithms. Sci Rep. 2019;9(1):1–11.

15. Cuff SM, Merola JP, Twohig JP, Eberl M, Gray WP. Toll-like receptor linked cytokine profiles in cerebrospinal fluid discriminate neurological infection from sterile inflammation. Brain Commun. 2020;2(2):1–15.

16. Rahbari NN, Weitz J, Hohenberger W, Heald RJ, Moran B, Ulrich A, et al. Definition and grading of anastomotic leakage following anterior resection of the rectum: A proposal by the International Study Group of Rectal Cancer. Surgery. 2010;147(3):339–51.

17. R Core Team. R: A language and environment for statistical computing. R Foundation for Statistical Computing [Internet]. Vienna, Austria; 2017. Available from: https://www.r-project.org/.

18. Cini C, Wolthuis A, D’Hoore A. Peritoneal fluid cytokines and matrix metalloproteinases as early markers of anastomotic leakage in colorectal anastomosis: A literature review and meta-analysis. Color Dis. 2013;15(9):1070–7.

19. Scheller J, Chalaris A, Schmidt-Arras D, Rose-John S. The pro- and anti-inflammatory properties of the cytokine interleukin-6. Biochim Biophys Acta - Mol Cell Res. 2011;1813(5):878–88.

20. Qi XY, Liu MX, Xu K, Gao P, Tan F, Yao ZD, et al. Peritoneal Cytokines as Early Biomarkers of Colorectal Anastomotic Leakage Following Surgery for Colorectal Cancer: A Meta-Analysis. Front Oncol. 2022;11(January):1–11.

21. Smith SR, Pockney P, Holmes R, Doig F, Attia J, Holliday E, et al. Biomarkers and anastomotic leakage in colorectal surgery: C-reactive protein trajectory is the gold standard. ANZ J Surg. 2018;88(5):440–4.

22. Sproston NR, Ashworth JJ. Role of C-reactive protein at sites of inflammation and infection. Front Immunol. 2018;9(APR):1–11.

23. Dong Q, Wright J. Expression of C-reactive protein by alveolar macrophages. J Immunol. 1996;156(12):4815–20.

24. Su’a B, Milne T, Jaung R, Jin JZ, Svirskis D, Bissett IP, et al. Detection of Anastomotic Leakage Following Elective Colonic Surgery: Results of the Prospective Biomarkers and Anastomotic Leakage (BALL) Study. J Surg Res [Internet]. 2022;273:85–92. Available from: https://doi.org/10.1016/j.jss.2021.12.019

25. Riese J, Schoolmann S, Beyer A, Denzel C, Hohenberger W, Haupt W. Production of IL-6 and MCP-1 by the human peritoneum in vivo during major abdominal surgery. Shock. 2000;14(2):91–4.

